# The Creation of a Multidomain Neighborhood Environmental Vulnerability Index Across an Urban Center

**DOI:** 10.1101/2022.12.14.22283064

**Authors:** Stephen P. Uong, Jiayi Zhou, Stephanie Lovinsky-Desir, Sandra S. Albrecht, Alexander Azan, Earle C. Chambers, Perry E. Sheffield, Azure Thompson, Joseph Wilson, Jennifer Woo Baidal, Jeanette A. Stingone

**Affiliations:** Columbia University, Mailman School of Public Health; Columbia University, Vagelos College of Physicians and Surgeons; NYU Langone Health, Department of Pediatrics at NYU Grossman School of Medicine; Albert Einstein College of Medicine, Department of Family and Social Medicine; Icahn School of Medicine at Mount Sinai, Department of Environmental Medicine and Public Health; SUNY Downstate Health Sciences University, School of Public Health

**Keywords:** area-level vulnerability, data integration, environmental justice, social determinants of health, deprivation

## Abstract

Compared to previous studies that have typically used a single summary score, we aimed to construct a multidomain neighborhood environmental vulnerability index (NEVI) to characterize the magnitude and variability of area-level factors with the potential to modify the health effects of environmental pollutants. Using the Toxicological Prioritization Index framework and data from the 2015-2019 U.S. Census American Community Survey and the 2020 CDC PLACES Project, we quantified census tract-level vulnerability overall and in 4 primary domains (demographic, economic, residential, and health status), 24 subdomains, and 54 distinct area-level features for New York City (NYC). Overall and domain-specific indices were calculated by summing standardized feature values within the subdomains and then aggregating and weighting subdomains within equally-weighted primary domains. In citywide comparisons, NEVI was correlated with both the Neighborhood Deprivation Index (r = 0.91) and the Social Vulnerability Index (r = 0.87) but provided additional information on features contributing to vulnerability. Vulnerability varied spatially across NYC, and hierarchical cluster analysis using subdomain scores revealed six patterns of vulnerability across domains: 1) low in all, 2) primarily low except residential, 3) medium in all, 4) high demographic, economic, and residential 5) high economic, residential, and health status, and 6) high demographic, economic and health status. Created using a tool that offers flexibility for theory-based construction, NEVI provided detailed metrics of vulnerability across domains that can inform targeted research and public health interventions aimed at reducing the health impacts from environmental exposures across an urban center.

## INTRODUCTION

Previous research has linked environmental pollutants, such as air pollution, toxic metals, and plastics, to various health outcomes, including cancer, neurodevelopmental impacts, asthma, and heart disease across various urban settings.^1–4^ Many prior studies conceptualized housing characteristics, poverty, health care access, and other neighborhood-level features individually as confounders or effect modifiers in evaluating the relationship between environmental pollutants and various health outcomes.^5–8^ In these studies, “neighborhoods” are spatial units where individuals reside that aim to capture unique group-level properties.^9^ These studies have demonstrated neighborhood-level factors can contribute to differences in environmental vulnerability, including greater likelihood of exposure to pollutants and greater susceptibility to health effects from environmental pollutants for individuals.

Composite neighborhood-level indices used to describe collective social and structural factors in previous work often measure a single domain of vulnerability or were not designed to focus on environmental vulnerability and flexible theory-based construction. For example, the commonly-used neighborhood deprivation index (NDI) that summarizes demographics, income, education, employment, and housing data have been used to measure neighborhood socioeconomic deprivation across different cities and health outcomes in environmental perinatal epidemiology studies.^10–14^ Similarly, the Centers for Disease Control and Prevention’s Social Vulnerability Index (SVI) summarizes several features to reflect neighborhood-level stressors relevant to community resilience to disasters or disease outbreaks.^15^ While there are newer efforts to further characterize neighborhood-level vulnerability such as the Climate and Economic Justice Tool, these tools often emphasize a single summary metric and provide limited ways to compare the relative magnitude or importance of individual features or domains that contribute to vulnerability between neighborhoods.^16^ Information about specific features or domains provides a more nuanced understanding of vulnerability that would inform targeted environmental justice efforts and urban planning policies by describing both the location of vulnerable neighborhoods and type of neighborhood-level characteristics that may be contributing vulnerability in those neighborhoods. Furthermore, indices like the NDI and SVI do not include other features that may increase environmental vulnerability, including pre-existing health conditions and behaviors that may increase vulnerability to health impacts from air pollution, heat stress, and climate change.^10,17–20^ Many indices have included neighborhood racial/ethnic composition as a component of vulnerability, although the measure does not contribute to environmental vulnerability but is rather a proxy for social and structural racist policies that led to segregation, disinvestment, and increased prevalence of other downstream outcomes that can increase vulnerability.^21,22^ Composite metrics designed to understand the social and structural drivers of environmental vulnerability should include the downstream effects of racism rather than using the neighborhood racial composition as a proxy for these effects. Finally, some indices like the SVI and the EPA/CDC Environmental Justice Index disaggregate vulnerability across domains but do not provide a framework for users to adapt the indices for their specific purpose, limiting their flexibility for theory-based construction.^23^

Previous research has used an approach called the Toxicological Prioritization Index (ToxPi)^24^ to combine multiple geospatial features into vulnerability indices across various spatial scales. In contrast to purely data-driven approaches commonly used in index construction,^10,14,25^ ToxPi allows researchers to subjectively retain features based on the specific research question of interest. Furthermore, ToxPi allows characterization of contributions to vulnerability across distinct domains, providing an ability to investigate whether neighborhoods may be experiencing different contributors to vulnerability and enabling tailored intervention approaches. For example, Bhandari et al applied ToxPi to the Houston-Galveston-Brazoria region to create a five-domain index designed to assist communities in developing plans to address health effects due to natural disasters and industrial activity.^26^ A similar framework was used by the National Institute of Environmental Health Science when constructing their COVID Vulnerability Index, to visualize national vulnerability to COVID-19^27^, again selecting domains based on their specific goals. Both prior approaches included environmental pollution as a component of their indices for overall vulnerability. While appropriate in some contexts, this limits the ability to investigate potential effect modification of the relationship between pollution and health outcomes by vulnerability.

The objective of this work was to apply the ToxPi approach to construct an adaptable multidomain neighborhood environmental vulnerability index (NEVI) in an urban center. We aimed to characterize the overall magnitude of and identify patterns in neighborhood environmental vulnerability within a large, demographically and socioeconomically diverse, and densely-populated urban area. This index was compared to previously-used indices to characterize the additional information gained from using a multidomain index and assess whether the index varies when constructed using downstream effects of racism rather than neighborhood racial/ethnic composition.

## METHODS

### Study Area

We focused on New York City (NYC), an urban center with intra-urban variability in a number of social and structural features that can potentially contribute to environmental vulnerability.^28^ To capture variation within NYC, we conceptualized our neighborhoods as census tracts, a relatively small spatial areal unit that could be aggregated into larger areas to correspond to other definitions of a neighborhood. In NYC, there are five distinct boroughs or counties (e.g. Bronx, Brooklyn, Manhattan, Queens, Staten Island), each with a unique development history that has shaped their current sociodemographic profile under their respective municipal jurisdictions. Thus, to maximize the policy and public health relevance of the NEVI, we describe its use in the context of these five boroughs.

Of the 2,167 census tracts in NYC, we excluded 51 tracts that had a population of less than twenty people and 30 tracts that had a population of at least twenty people but had missing data for at least one feature. The majority of these 30 census tracts were in non-residential areas, such as construction sites, parks, and areas with institutions like prisons or universities. As a result, we ultimately included 2,086 census tracts in the NEVI. This analysis is part of a research protocol reviewed and approved by the Columbia University Institutional Review Board.

### Construction of Neighborhood Environmental Vulnerability Index

To construct the NEVI, we used data from the 2015-2019 U.S. Census American Community Survey (ACS) 5-year estimates and the 2020 data release from the Centers for Disease Control and Prevention (CDC) PLACES Project. ToxPi is a tool to integrate and visualize data across multiple domains.^29^ Subject-matter knowledge is used throughout the process, including the selection of features, grouping the features into subdomains (and subdomains into domains), and weighting of all components to generate the overall index. To select our domains, subdomains, and distinct area-level features, we conducted a literature search of previous research on social and structural drivers of vulnerability to environmental pollution.^5,6,17,19,30–32^ Furthermore, we reviewed previously published vulnerability indices to compile and adapt domains from previous applications of ToxPi for our purpose.^10,25–27,33^ For example, the HGBEnviroScreen tool included a social vulnerability domain that we disaggregated into demographic and economic domains and three other domains that measured environmental risks and exposures that we did not include to allow for later potential effect modification analyses by those environmental measures.^26^ Guided by the literature and subject-matter knowledge of the research team, we selected 4 primary domains composed of 24 subdomains and 54 distinct area-level features that were hypothesized to contribute to potential environmental vulnerability. The four primary domains were 1) demographics, 2) economic indicators (“economic”), 3) residential characteristics and density (“residential”), and 4) health behavior, outcomes, preventative practices, and access (“health status”) (Table 1). Features within the demographics, economic, and residential domains were from the U.S. Census American Community Survey, while features within the primary domain of health status were drawn from the CDC PLACES Project (model-based estimates).^34,35^

**Table 1.**
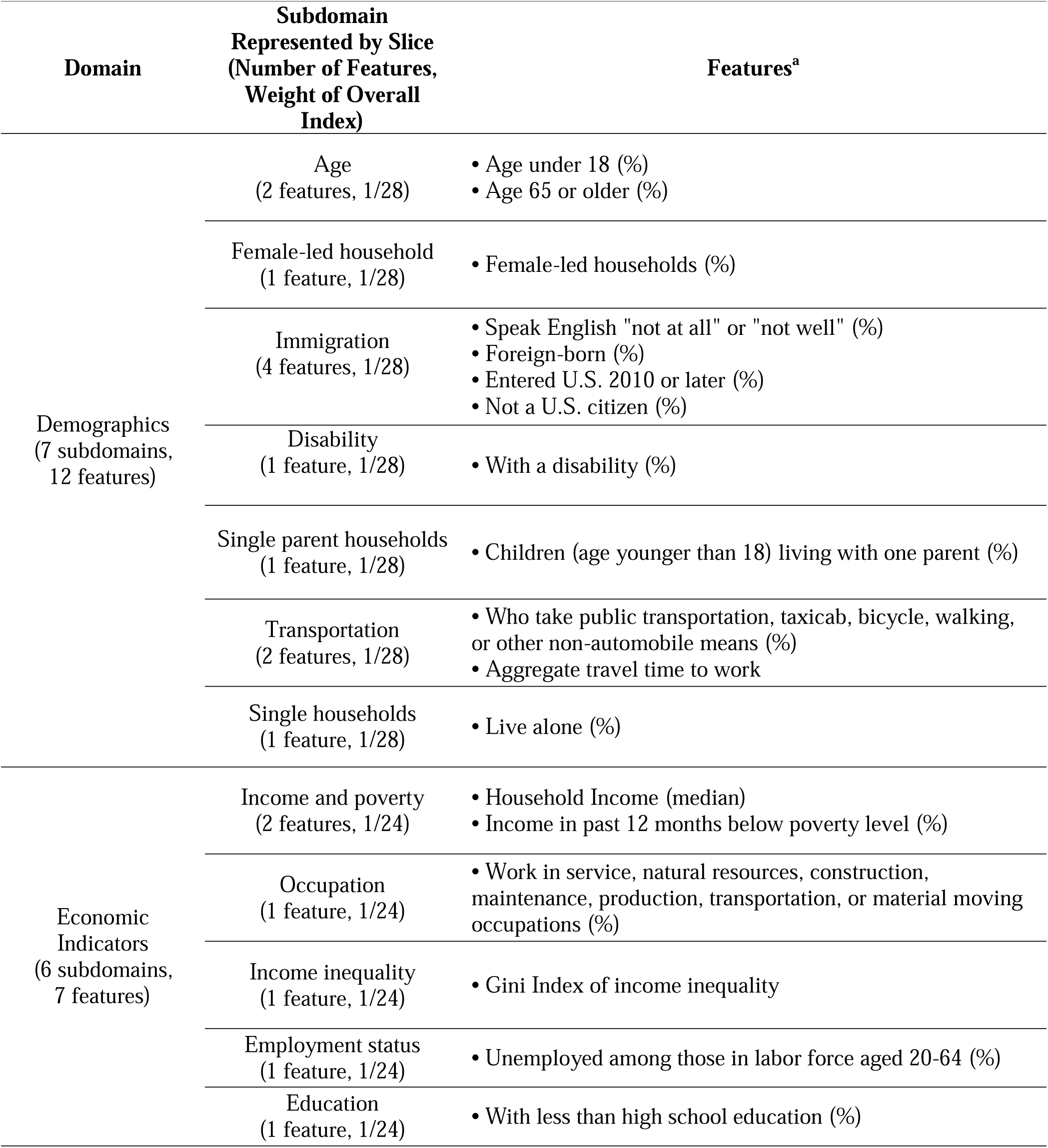
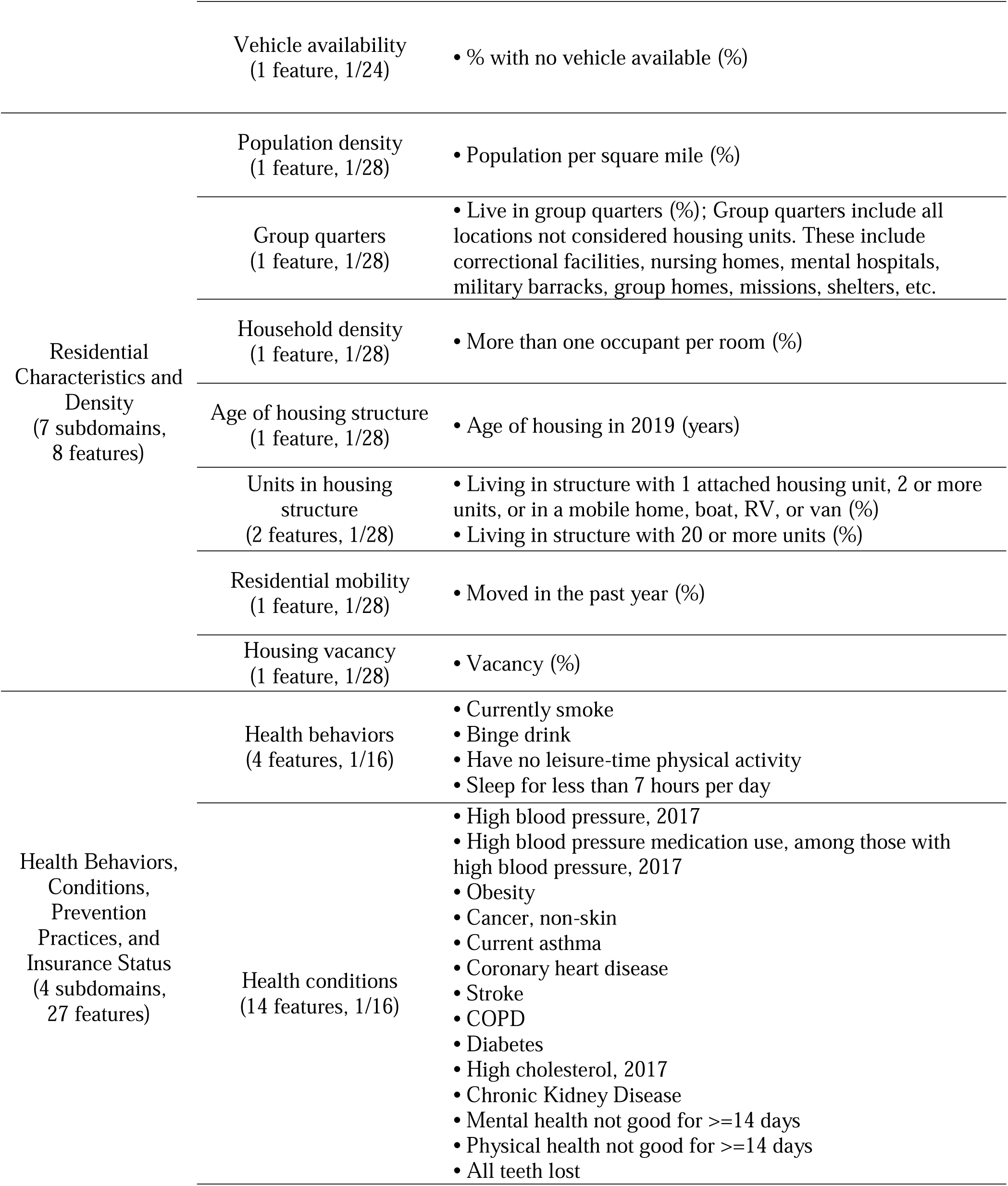
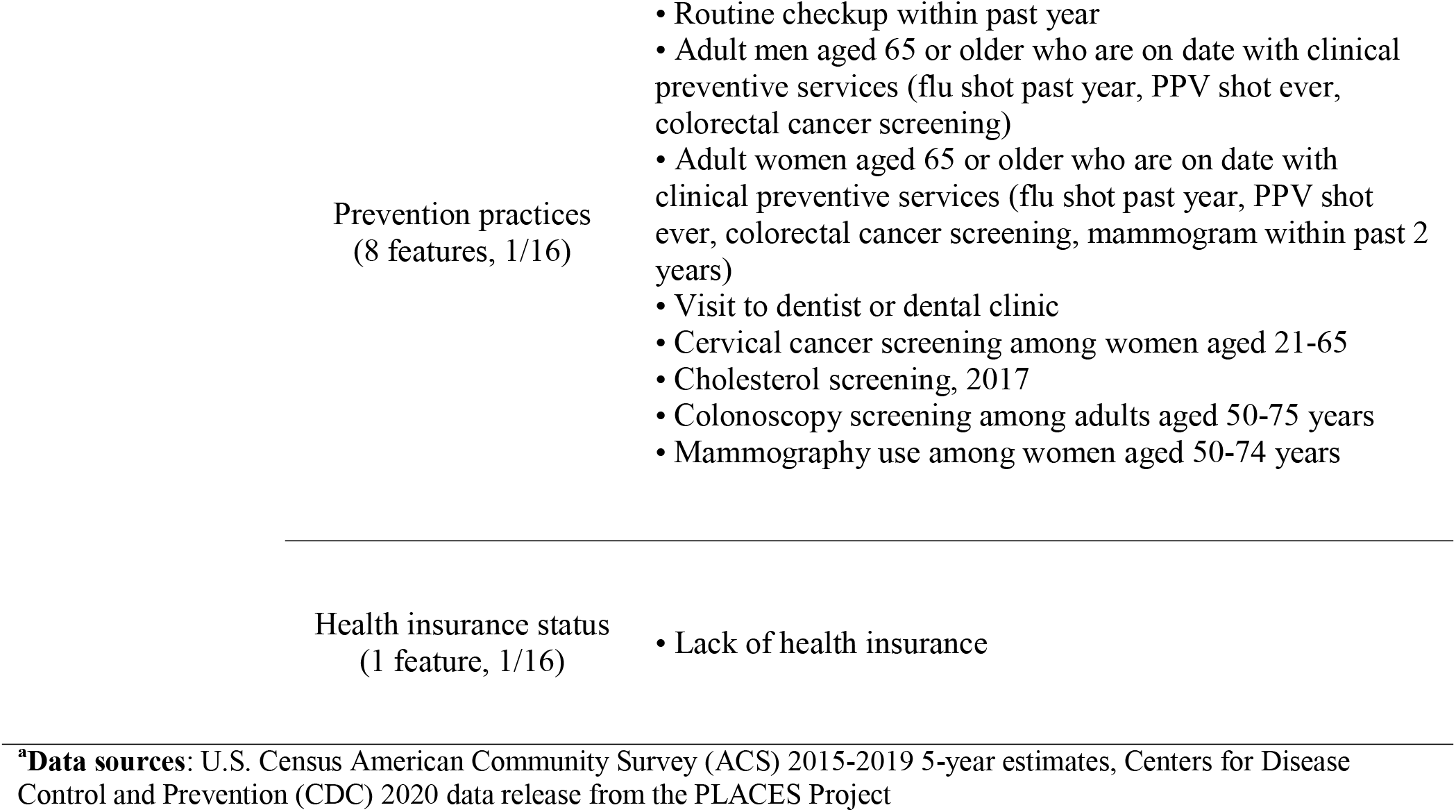
Features Included in the Neighborhood Environmental Vulnerability Index.

The demographic domain consisted of 7 subdomains: age distribution, female-led households, nativity, disability, single parent households, transportation behaviors, and single households. For our primary analyses, we did not include racial and ethnic composition because it is systemic racism manifested as racist policies rather than racial or ethnic identity that lead to greater social and economic stressors and environmental vulnerability.^36^ Instead, we only included stressors in the index that reflect both individual-level aggregates and neighborhood-level characteristics. We conducted a sensitivity analysis to see if the NEVI would vary upon the inclusion of race and ethnicity composition. The economic domain included 6 subdomains: income and poverty status, occupation, income inequality, employment status, education, and vehicle availability. The residential domain included 7 subdomains: population density, group quarters, household density, age of housing structure, number of units in housing structure, residential mobility, and housing vacancy. Finally, the health status domain included 4 subdomains: health behaviors, health conditions, prevention practices, and health insurance status.

We used the ToxPi Graphical User Interface to calculate the overall NEVI and domain-specific scores, conceptualizing each slice as a subdomain within our four domains.^37^ We first standardized each feature with z-scores. Next, the software summed the values across the features within each subdomain before transforming the summed values by subtracting the minimum value and dividing by the range of the values.^38^ The resulting subdomain scores were then multiplied by the weights (specified in Table 1) to calculate the overall NEVI.^38^ While software calculates the overall NEVI and subdomain-specific scores, we manually calculated domain-specific scores by averaging the subdomain-specific scores within each of the four domains. Because ToxPi converts negative values to zero, we re-centered features so that the minimum value would equal zero, resulting in values that were zero or greater across all features. We coded features to ensure that greater (i.e., more positive) values would indicate greater vulnerability. The final NEVI overall and domain-specific scores ranged from 0 to 1.

### Comparison to Existing Indices: NDI and SVI

For comparison, we constructed an adapted Neighborhood Deprivation Index (NDI) originally developed by Messer et al. from the 2015-2019 U.S. Census ACS and used the Social Vulnerability Index (SVI) prepared by CDC.^39^ To construct a tract-level NDI that reflects heterogeneity across spatial strata of NYC, we adapted methods from both Messer et al. and Shmool et al (described in greater detail in the supplementary material).^14,39^ Briefly, after conducting successive principal component analyses (PCA), 7 features were retained from a pool of candidate indicators of area-level deprivation: percent population with at least Bachelor’s level education, percent unemployed, adults in management or professional occupations, households in poverty (< 200% Federal Poverty Line), families with annual income less than $35,000, households with public assistance income, and proportion of residents who were persons of color (Supplemental Table S1). These seven features were included in a final PCA and the resulting first unrotated principal component was used as the tract-level NDI.

### Descriptive and Statistical Analyses

We calculated and described the distributions of the NEVI (overall, by domain, and by borough), NDI, and SVI. Next, we mapped both the overall index and the domain-specific scores. To compare the NEVI to the NDI and SVI, we visualized distributions of quartiles of the indices in adjacent maps and calculated Spearman correlation coefficients between the NEVI and NDI/SVI across domain and borough. To identify common patterns in the NEVI subdomains across census-tracts, we conducted a hierarchical clustering analysis with complete linkage and selected the optimal number of clusters using the Gap-statistic.^40^ The composition of resulting clusters was compared using heat maps of the median subdomain scores, standardized by feature to compare relative scores across clusters. As a sensitivity analysis, all analyses were repeated after including racial and ethnic composition features within the demographic domain of the NEVI. Specifically, we included 4 features describing the proportion of residents who identified as Hispanic/Latino of any race, Black non-Hispanic/Latino, Asian non-Hispanic/Latino, and other non-white Race non-Hispanic/Latino. Finally, we summarized overall and domain-specific vulnerability by high racial and ethnic composition in a given neighborhood, defined as having a racial and ethnic composition higher than the citywide median proportion. All data preprocessing and analyses were completed in R version 4.0.2. We used the *tidycensus* package to download data from the U.S. Census,^41^ the *nycgeo* package to download shapefiles of NYC census tracts included in our choropleth maps,^42^ and the *psych* package to perform PCA.^43^ All programming code and data are available on Github jstingone/nevi.

## RESULTS

### Overall NEVI and Domain-Specific Scores

Figure 1 displays the distribution of the overall NEVI along with the domain-specific scores in separate maps. Numeric estimates of the distribution of the overall index and domain-specific scores for all census tracts are provided in Supplemental Table S2.

**Figure 1.**
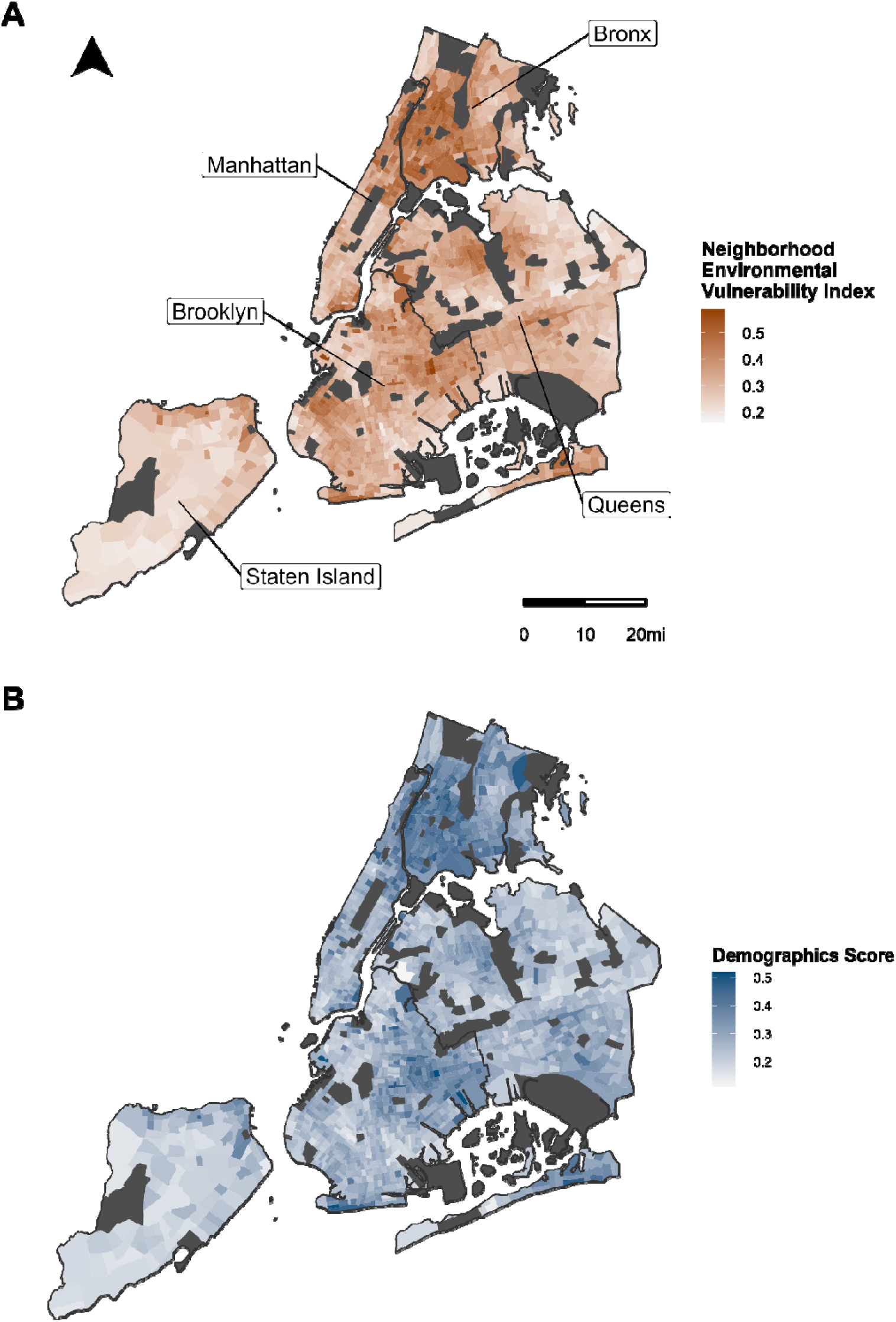
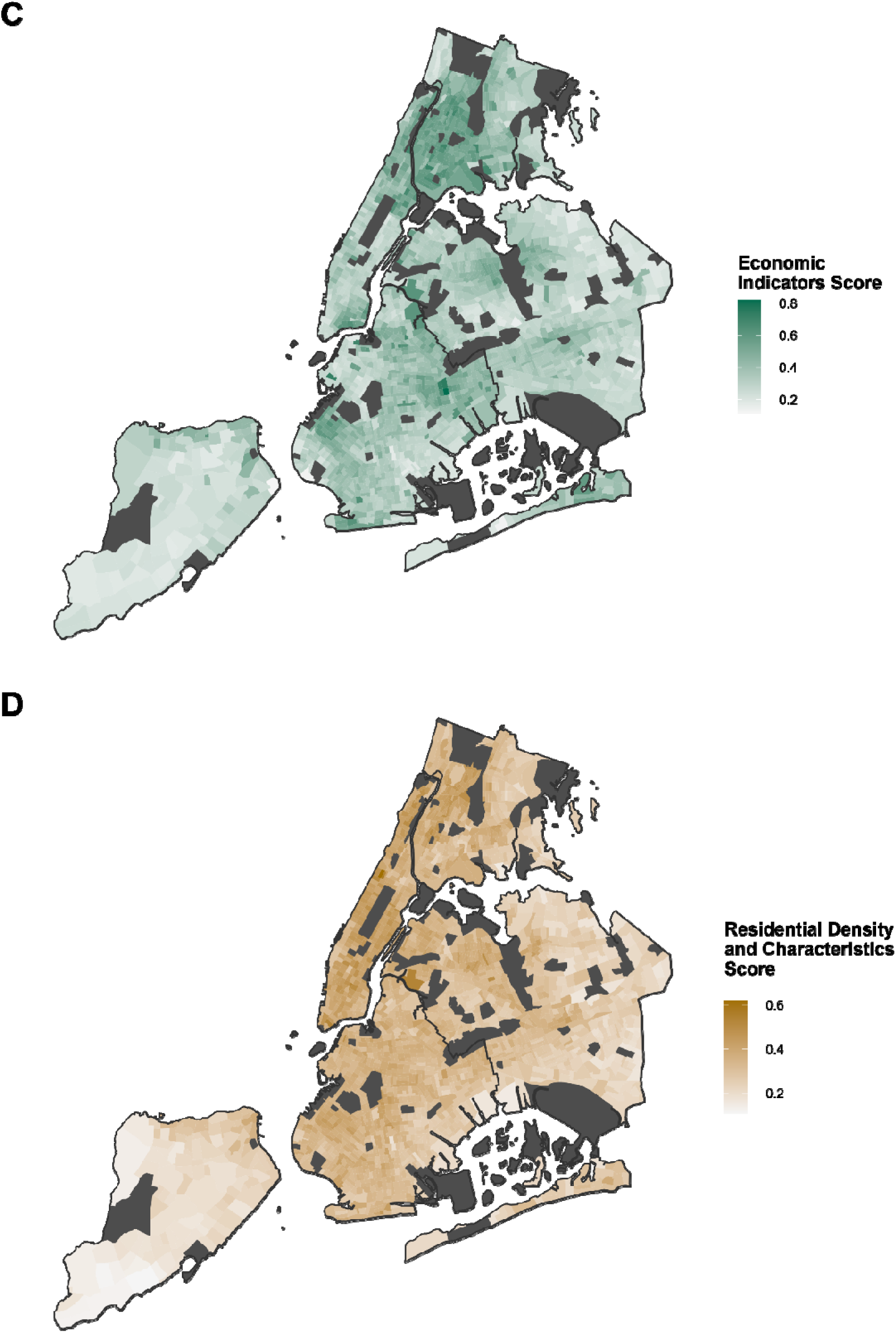
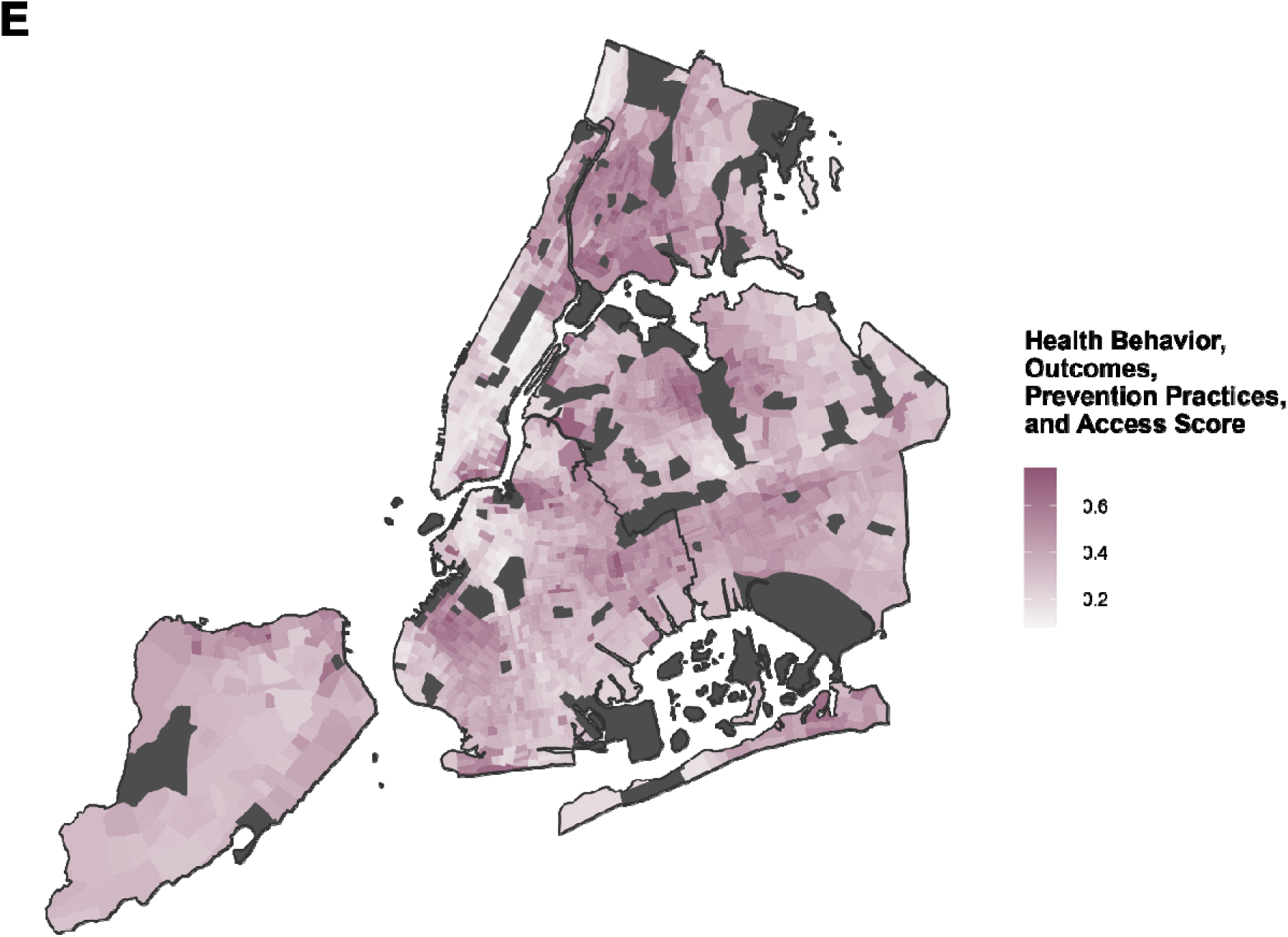
Maps of Neighborhood Environmental Vulnerability Index: Overall Index and Domain-Specific Scores Across New York City by Census Tract, 2015-2019. The maps display the distribution of the (A) overall Neighborhood Environmental Vulnerability Index across New York City along its (B-E) domain-specific scores. Areas that were excluded due to low population counts or missing features are shown in dark gray. Data Sources: U.S. Census American Community Survey 2015-2019 5-Year Estimates and Centers for Disease Control and Prevention PLACES Project 2020 Release

As shown in Figure 1 A-D, the NEVI had variability in both overall magnitude and distribution of the domains when comparing the index across boroughs (Supplemental Table S2). The Bronx, an outer borough with mid-range population density and the lowest median income in NYC, had the highest median index (0.43, IQR = 0.35-0.48), while Staten Island, the borough with the lowest population-density and second highest median income, had the lowest median index (0.25, IQR = 0.23-0.30). Consistent with the overall index, median domain scores were the highest in the Bronx for the demographics (0.38, IQR = 0.32-0.41), economic (0.48, IQR = 0.37-0.57), and health status (0.49, IQR = 0.35-0.56) domains and the lowest in Staten Island for the demographics (0.21, IQR = 0.19-0.26), economic (0.25, IQR = 0.22-0.30), and residential (0.21, IQR = 0.18-0.25) domains (Supplemental Table S2, Supplemental Figure S1). However, the median residential domain score was the highest (0.41, IQR = 0.37-0.44) and median health status domain score was the lowest (0.24, IQR = 0.16-0.42) in Manhattan, the borough with the highest population-density but also the highest median income.

### Comparisons Between NEVI, NDI, and SVI

Overall, there were strong, positive correlations between NEVI and NDI (r = 0.91, p < 0.001) and NEVI with SVI (r = 0.87, p < 0.001). However, there were some notable differences when visually comparing the distribution patterns of these three indices by borough, including less agreement between the indices in Queens, the most ethnically-diverse borough^44^ (Figure 2). Correlations between the overall NEVI and NDI/SDI were lower in Queens than other areas, especially for the NDI (Supplemental Figure S2). The NDI did not explain as much variance in Queens as in other boroughs. Correlations between the domain-specific scores and the NDI and SVI were high, except between the NEVI residential score which consistently showed lower correlations with both the NDI and the SVI across boroughs.

**Figure 2.**
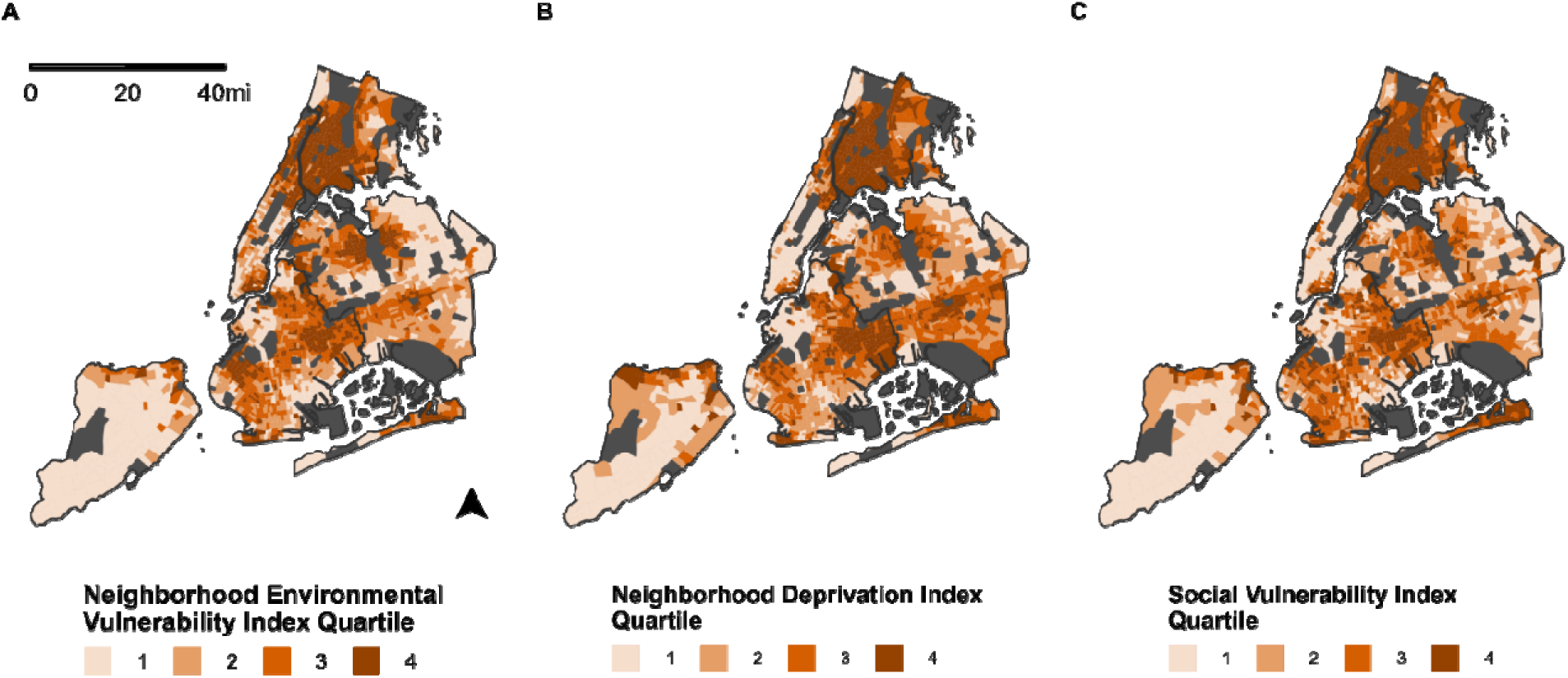
Maps of Neighborhood Environmental Vulnerability Index, Neighborhood Deprivation Index, and Social Vulnerability Index Across New York City by Census Tract, 2015-2019. The maps display the distribution of the (A) overall Neighborhood Environmental Vulnerability Index, (B) the Neighborhood Deprivation Index, and (C) the Social Vulnerability Index across New York City. Areas that were excluded due to low population counts or missing features are shown in dark gray. Data Sources: U.S. Census American Community Survey 2015-2019 5-Year Estimates, Centers for Disease Control and Prevention PLACES Project 2020 Release, and Centers for Disease Control and Prevention/Agency for Toxic Substances and Disease Registry 2018 Social Vulnerability index

### Vulnerability Profiles of Census Tracts

As an example of how domain-specific features contribute differently to NEVI and support a targeted and adaptable public health approach, in Figure 3, we show vulnerability profiles for two sample census tracts in NYC with similar overall NEVI scores (0.239): Census Tract 671 and Census Tract 519. Despite having similar overall scores, the patterns across subdomain scores are different with greater economic vulnerability in Census Tract 519 (0.64 vs 0.23 economic sub-score) and slightly greater scores in residential and health-related features in Census Tract 671.

**Figure 3.**
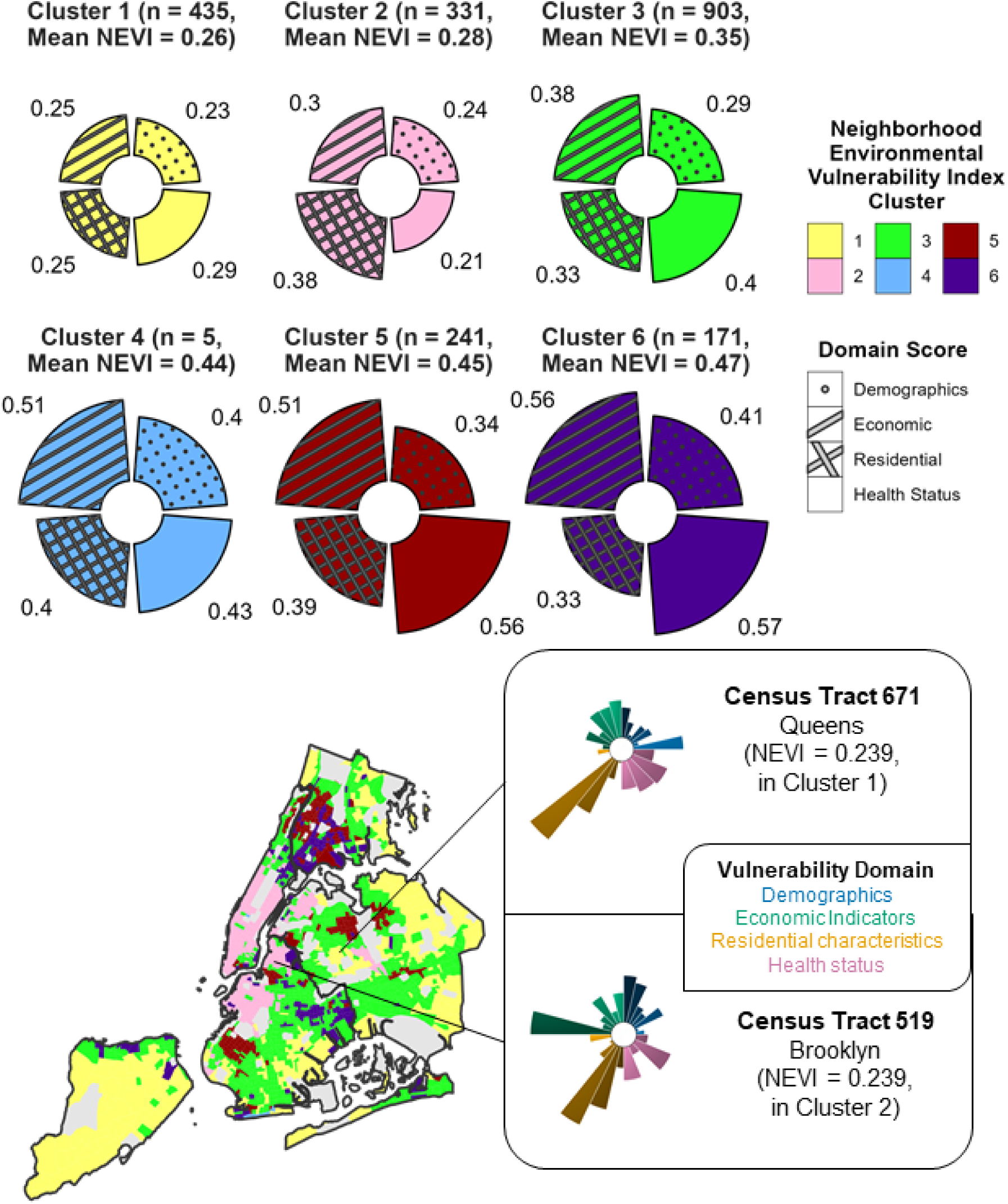
Map of Clusters with Example Tract Vulnerability Profiles for the Neighborhood Environmental Vulnerability Index across New York City, 2015-2019. The vulnerability profiles for the six clusters (on top) and two census tracts are shown: Census Tract 671 in Queens and Census Tract 519 in Brooklyn (on bottom right). The different colors represent different clusters in the cluster vulnerability profiles, and the different colors in the Census tract vulnerability profiles represent different domains. Within each domain of the Census tract vulnerability profiles, the different shades of each slice represent various subdomains, with larger slices representing greater vulnerability. The map (bottom left) shows the distribution of the NEVI clusters across NYC, with areas that were excluded due to low population counts or missing features shown in dark gray. Data Sources: U.S. Census American Community Survey 2015-2019 5-Year Estimates and Centers for Disease Control and Prevention PLACES Project 2020 Release.

### Vulnerability Profiles of NEVI Clusters

Our clustering analysis revealed 6 distinct patterns in NEVI domain-scores across census-tracts (Figure 3), specifically two low vulnerability clusters, one medium vulnerability cluster and three high vulnerability clusters: Cluster 1) low across all domains (median NEVI = 0.26, n = 435); Cluster 2) primarily low but greater residential vulnerability and very low health vulnerability (median NEVI = 0.28, n = 331); Cluster 3) medium vulnerability across all domains (median NEVI = 0.35, n = 903); Cluster 4) high vulnerability across demographic, residential and economic domains (median NEVI = 0.44, n = 5); Cluster 5) high vulnerability across economic, residential and health status domains (median NEVI = 0.45, n = 241); and Cluster 6) high vulnerability across demographic, economic and health status domains (median NEVI = 0.47, n = 171). Figure 4 provides a heatmap to enable greater visualization of differences in specific subdomains across clusters. For example, the low vulnerability Cluster 2 had consistently lower health status vulnerability scores than Cluster 1 but tended to have higher scores in age of housing structure, units in housing structure, residential mobility, and vacancy within the residential domain. Additionally, the high vulnerability Cluster 6 has consistently lower residential vulnerability than the other high vulnerability clusters, except for the location of group quarters but higher vulnerability related to employment status and single-parent or female-led households.

**Figure 4.**
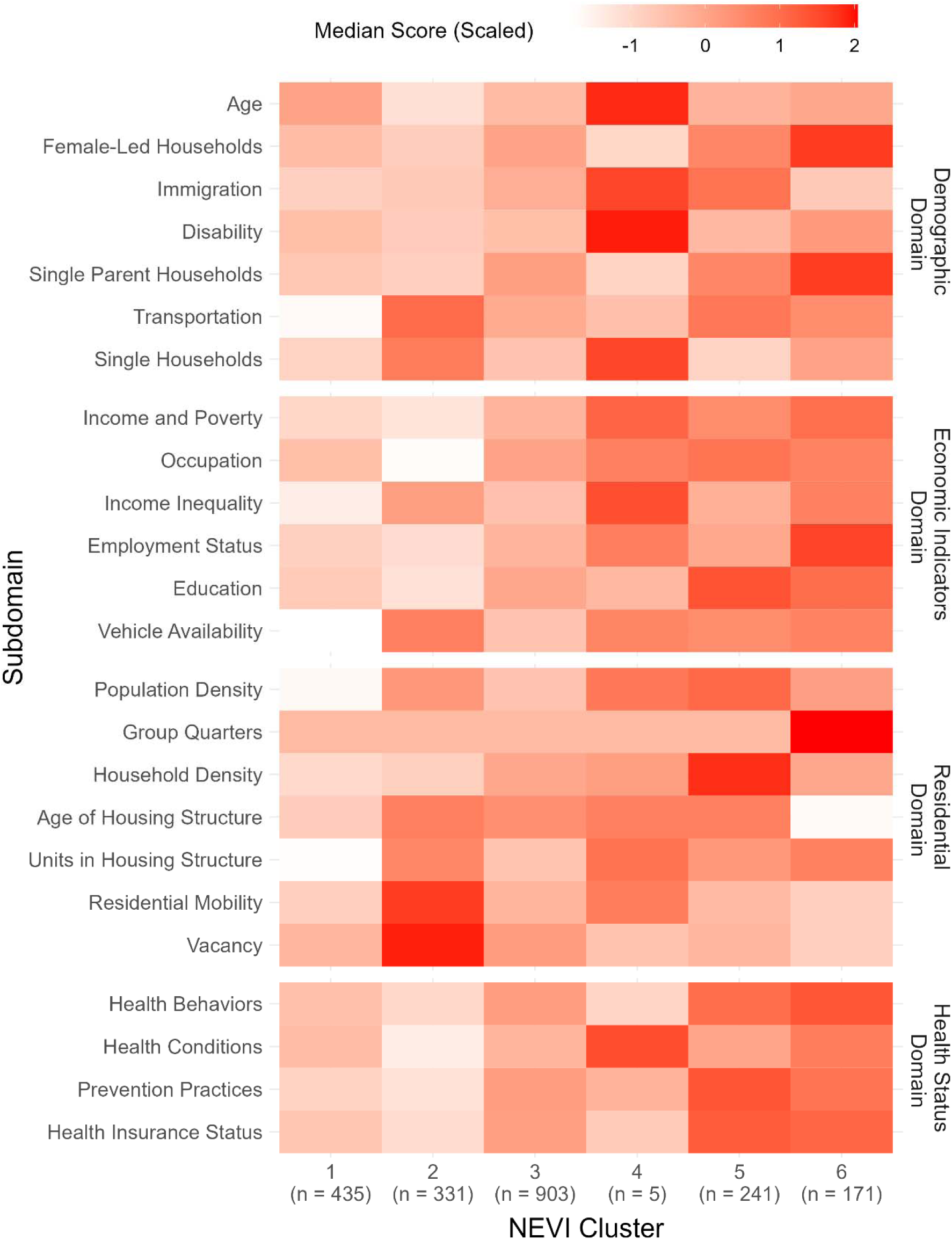
Heatmap of Median Subdomain Scores by Neighborhood Environmental Vulnerability Index Cluster. The median subdomain scores were standardized by feature to emphasize the relative magnitude of the subdomain scores across clusters

**Figure 5.**
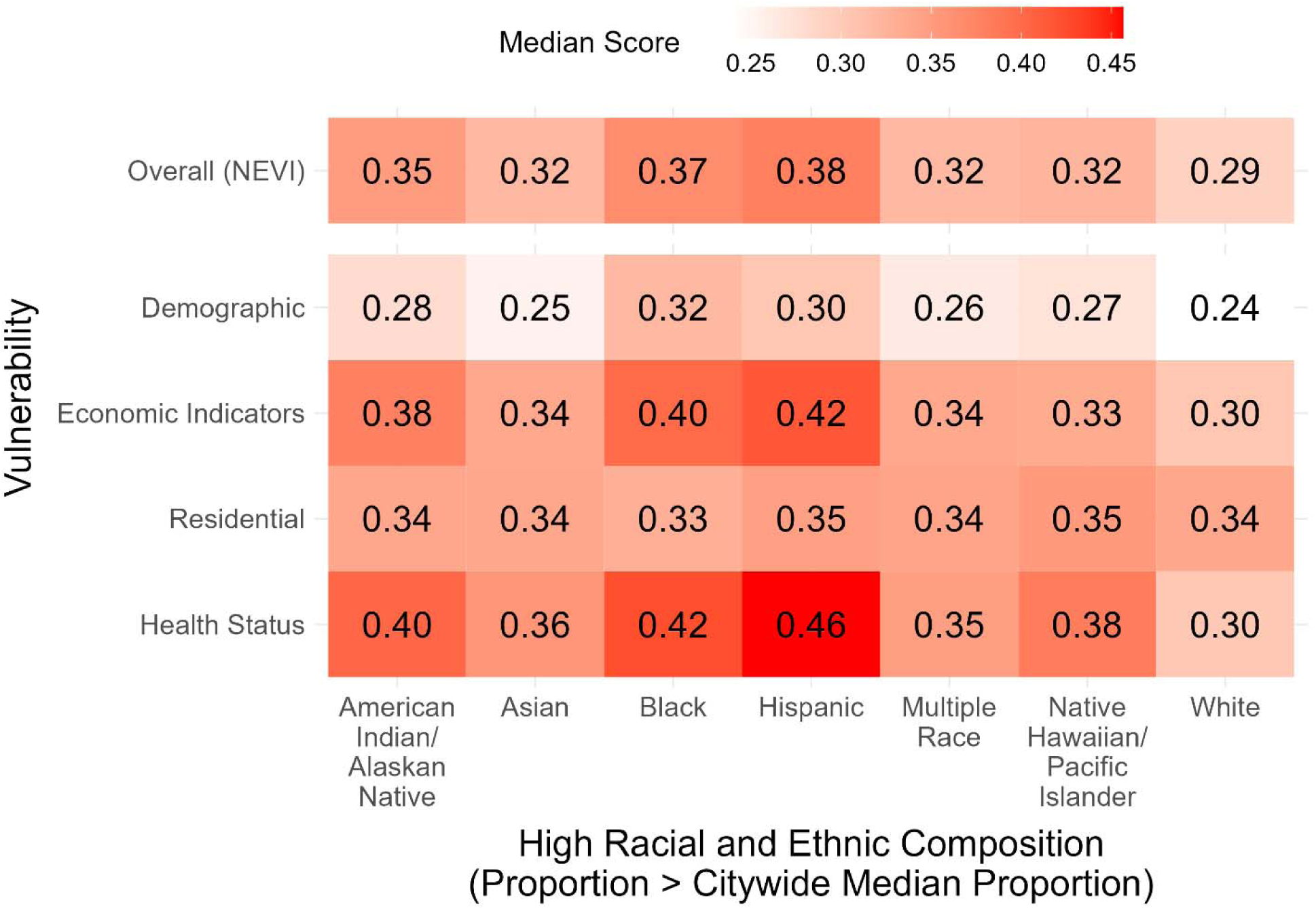
Median Overall and Domain Scores by High Racial and Ethnic Composition. The median NEVI and domain-specific scores are shown by high racial and ethnic composition across neighborhoods, defined as having a racial and ethnic composition higher than the citywide median proportion. A given neighborhood may have a high composition across multiple racial and ethnic groups (e.g., a neighborhood with both 1) % Hispanic higher than the citywide median % Hispanic and 2) % Native Hawaiian/Pacific Islander higher than the citywide median % Native Hawaiian/Pacific Islander).

### Sensitivity Analyses by Race and Ethnicity

The NEVI did not change considerably when including racial and ethnic composition of census tracts as features. Supplemental Figure S3 illustrates that the mean values and distributions for the overall NEVI did not? change in NYC as a whole or by borough when including racial and ethnic composition. The Demographic domain scores changed slightly, particularly in Queens where inclusion of race/ethnicity features caused slightly higher mean domain scores. Cluster patterns changed slightly when including race/ethnicity features in the NEVI (Supplemental Figure S4). For example, some medium vulnerability census-tracts were regrouped with the low vulnerability clusters, leading to more low-vulnerability areas. Some previously high vulnerability census-tracts were reclassified as a medium vulnerability cluster. This caused small changes in the overall vulnerability scores of clusters, but the majority of census-tracts remained within the same cluster.

In comparisons of vulnerability across race and ethnicity, overall vulnerability was lowest in White (neighborhood composition higher than the citywide median proportion) neighborhoods (median NEVI = 0.29), moderate in Asian, Multiple Race, and Native Hawaiian/Pacific Islander neighborhoods (median NEVI = 0.32), and highest in American Indian/Alaskan Native, Black, and Hispanic neighborhoods (median NEVI = 0.35, 0.37, and 0.38, respectively). When disaggregating by domain, residential vulnerability was similar across racial and ethnic groups, and economic and health status vulnerability was especially high among American Indian/Alaskan Native, Black, and Hispanic neighborhoods.

## DISCUSSION

The NEVI characterized neighborhood environmental vulnerability across NYC and quantified contributions to vulnerability across four domains: 1) demographics, 2) economic indicators, 3) residential characteristics, and 4) health status. There was general agreement between the overall NEVI and previously developed indices for deprivation (NDI) and social vulnerability (SVI). However, the NEVI offered additional benefits, including its adaptable construction and additional information about the types of vulnerability across a geographic area. Together, these can inform efforts to enhance subsequent environmental justice research and interventions aimed at reducing vulnerability to environmental pollutant exposures in neighborhoods at the hyperlocal scale.

Our index characterized potential contributions towards vulnerability from different domains, in contrast to previous indices that only provide an overall score for vulnerability or deprivation.^10,45^ Prior studies that used a single measurement for deprivation were limited in their ability to describe which aspect of the neighborhood environment were related to their results.^46,47^ Knowing which domains contribute to higher neighborhood vulnerabilities would facilitate adaptable population health planning and research to allocate certain types of resources to targeted neighborhoods even if the domain or specific features are not themselves modifiable, such as tailoring interventions to specific vulnerable demographic groups in a given neighborhood. Our clustering analysis revealed six primary patterns of potential vulnerability, which differentiated boroughs in more detail than conveyed by the absolute magnitude. For example, clusters 1 and 2 both had low overall vulnerability, but cluster 2 had even lower vulnerability due to existing health conditions but greater vulnerability associated with residential characteristics. This could point to different routes of intervention to reduce and/or ameliorate related health impacts of environmental pollution. For example, when designing an intervention to reduce the health effects of air pollution, one may target better chronic disease management in Cluster 1 while focusing on improved ventilation in the older housing stock of Cluster 2.

Notably, we excluded measures of racial and/or ethnic composition in the construction of our final index. Communities of color disproportionately experience worse health outcomes resulting from exposure to environmental pollutants and greater amounts of social and economic stressors across the US. For example, racial segregation along with historic racist land use policies such as red-lining have led to concentrated poverty and structural disinvestment in racially minoritized and low income urban neighborhoods that could increase their vulnerability to the health impacts of environmental pollutants.^31,32,48–51^ Rather than include the neighborhood racial and ethnic composition as a feature in the NEVI, we included the downstream effects of racism, such as income inequality and degree of chronic disease burden. Our sensitivity analyses revealed that there was little difference in overall NEVI scores based on the inclusion of racial/ethnic composition, likely due to the inclusion of socioeconomic, residential and health-related outcomes that are highly correlated with race through the impacts of racism. A previous analysis of the Climate and Economic Justice Screening Tool, developed by the White House, also found similarity in community rankings regardless of whether racial and ethnic demographics were included.^52^ However, other contexts seeking to use an index like the NEVI may want to include racial and/or ethnic composition as proxies for other measures of racism and structural inequities that remain unmeasured. This separation of race and ethnic composition from NEVI allowed us not only to identify higher overall vulnerability in communities of color but also that the variation in vulnerability was driven by differences in economic indicators and health status, reflective of racial and ethnic disparities seen in the US. We recommend that future studies further validate the NEVI across other exposures and in causal analyses with health outcomes.

The NEVI and NDI/SVI were generally similar across boroughs and domains, except the NEVI residential domain (i.e., lower correlations), likely because no or fewer housing-related features linked to environmental hazards was included in the NDI/SVI.^5,53,54^ Overall, NEVI had a lower correlation with NDI/SVI in Queens and the NDI explained less total variation in Queens compared to other NYC boroughs, possibly because Queens is the most demographically and socioeconomically diverse borough in NYC.^44,55^ A limitation of the NEVI is the need to exclude some Census tracts due to low population or missing data for at least one feature, limiting its potential applicability in those areas. These areas are composed of institutions like prisons, in which people may have increased environmental vulnerability.^56,57^ Additionally, we have currently only used the NEVI in one large, demographically and socioeconomically diverse, and densely populated urban center, potentially limiting the generalizability of our observed patterns in vulnerability to other urban areas. However, the adaptable NEVI creation process would facilitate the creation of the index in other urban areas to ascertain their distinct patterns of potential vulnerability.

The NEVI provides several advantages that increases its interpretability and utility. First, we were able to characterize the magnitude of potential environmental vulnerability, quantify the contributions from various domains, and identify patterns of vulnerability across an urban area. These provide additional interpretability that would better inform public health planning and additional domain-specific scores to evaluate in research studies incorporating vulnerability measures. Next, ToxPi enables the use of background knowledge to inform feature selection and weighting approaches, unlike indices that use data-driven methods to retain features and may drop features of theoretical importance but are highly correlated with other features. Being able to choose features and specify weights for contributions to vulnerability allows for a hybrid approach in which more hypothesis-driven information may be incorporated in the index. The process of customizing the NEVI in choosing features and specifying weights to fit specific hypotheses is transparent, facilitating discussion and critique. For example, we selected four domains that were hypothesized to reflect distinct area-level characteristics that could contribute to environmental vulnerability, distinct from the concentrations of environmental pollution that may exist in a neighborhood that might be included in other indices to identify areas most at-risk of specific health outcomes. In contrast to other indices like the SVI that includes four pre-set themes, the use of ToxPi enables the addition of domains and/or features in a clear manner, that promotes flexibility for the investigator and transparency for communities and stakeholders seeking to interpret the vulnerability index.

## Conclusion

We developed a neighborhood-level index to measure vulnerability to health impacts from environmental pollution for New York City and found that our NEVI was generally consistent with previously-developed deprivation and vulnerability scores. However, the NEVI was additionally able to characterize contributions to vulnerability across multiple domains, providing greater insight into intra-urban variation in vulnerability. Specifically, the customization option of this index-building approach allowed theory-based analysis of specific features/domain contribution to the index score (as we explored with racial composition). This metric can be used to inform targeted public and environmental health research and practice at the hyperlocal scale and improve our understanding of the impact of environmental exposures on communities with varying levels of vulnerability.

## Data Availability

All data and code produced are available online.

https://github.com/jstingone/nevi

## ACKNOWLEDGEMENTS

The authors would like to thank Denise Nguyen and Tamia E. Wellington for their help with preliminary analyses and members of Health Data for New York City (HD4NYC) for their feedback on the creation of the NEVI and this manuscript.

