## Supplemental Material for "The Creation of a Multidomain Neighborhood Environmental Vulnerability Index Across an Urban Center"

**SUPPLEMENTARY INFORMATION**

**Supplemental Figure 1. Distributions of the Neighborhood Environmental Vulnerability Index, Neighborhood Deprivation Index, and Social Vulnerability Index by Boroughs in New York City**

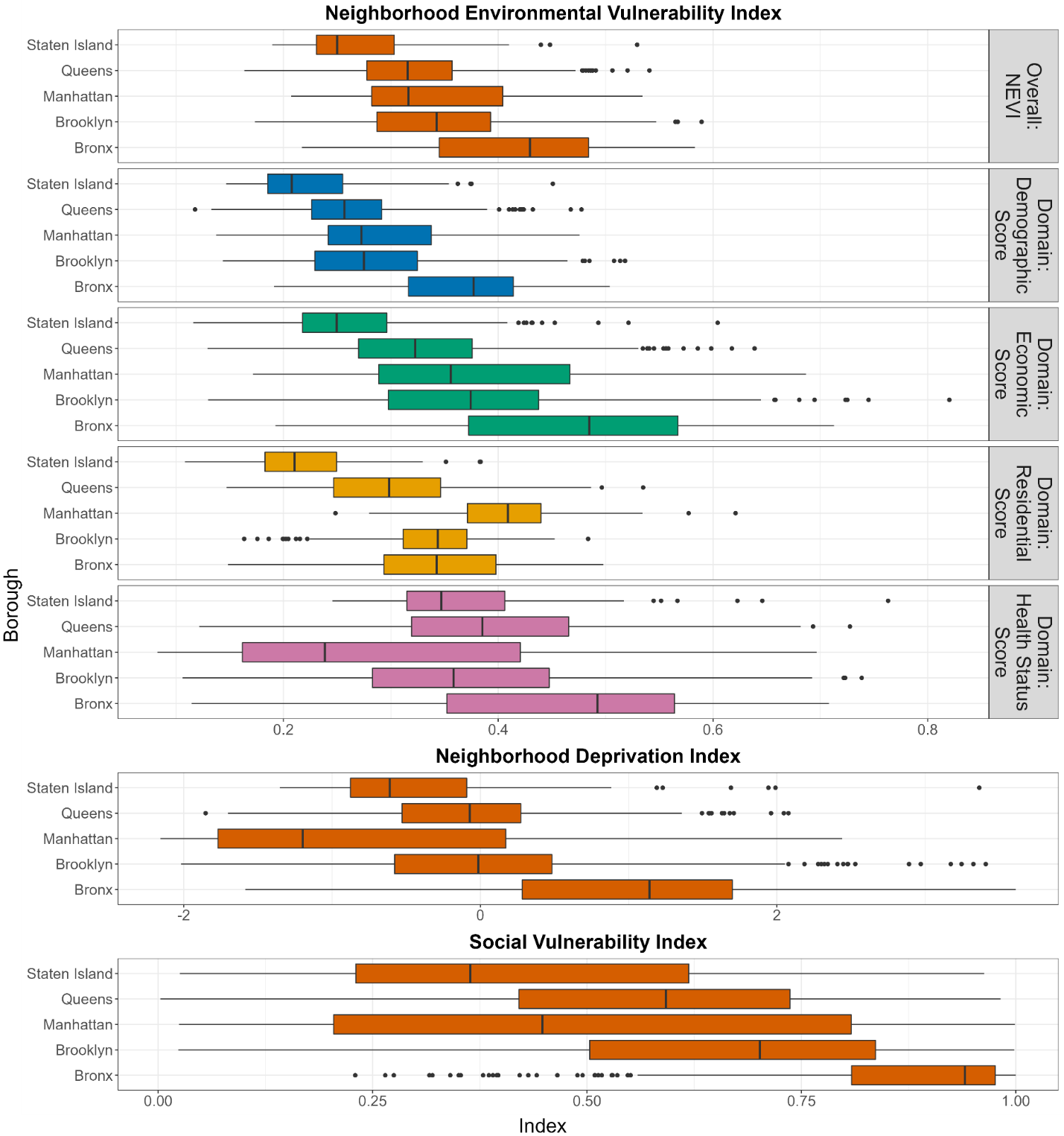

The boxplots above display the median, first and third quartile represented by the lower and upper hinges, first or third quartile ± 1.5*IQR (interquartile range) represented by the lower and upper whiskers, and outliers beyond the whiskers represented by the dots.

**Supplemental Figure 2A. Correlations Between Neighborhood Environmental Vulnerability Overall Index and Domain-Specific Scores with Neighborhood Deprivation Index by New York City Boroughs, 2015-2019**

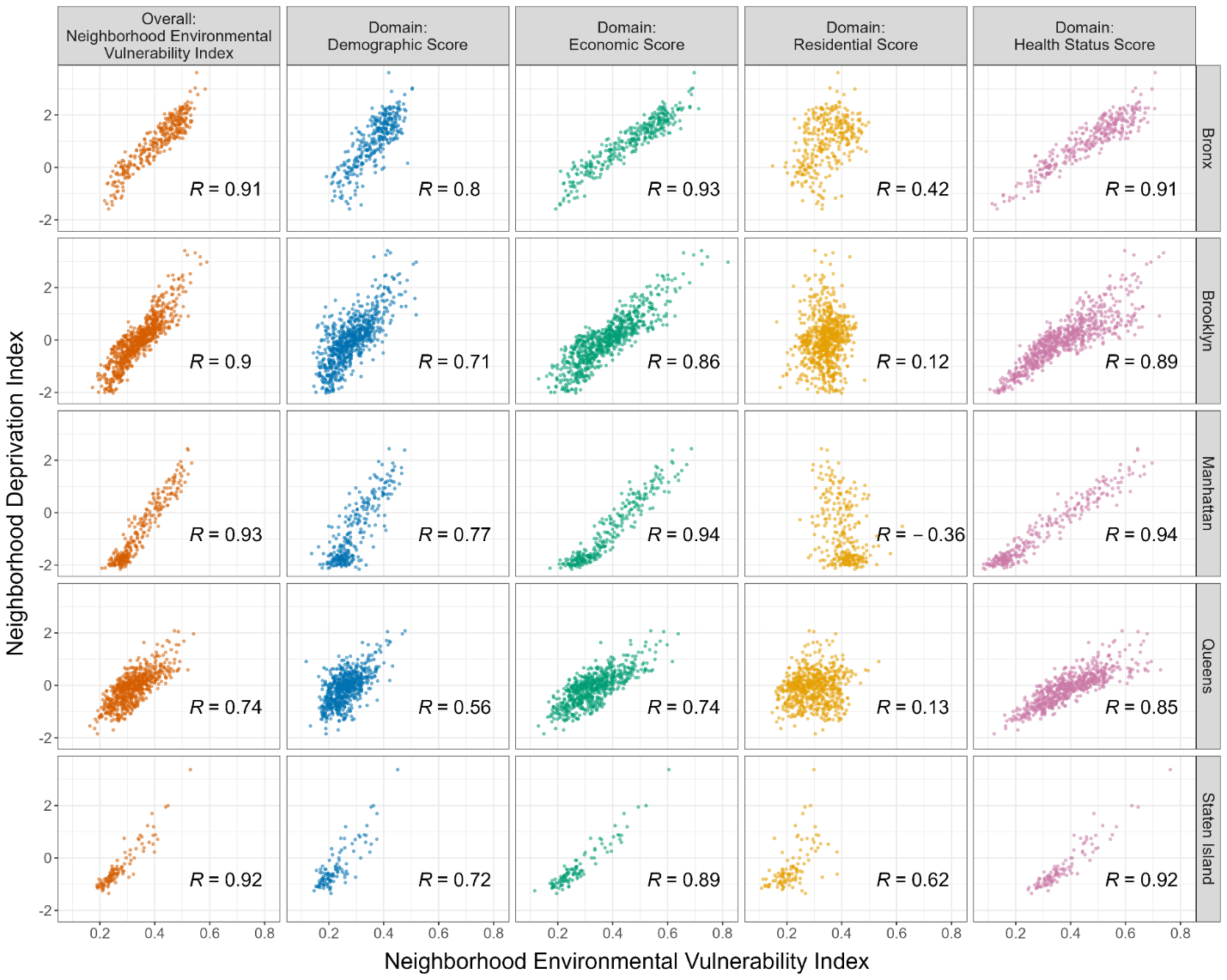

Each point represents the Neighborhood Environmental Vulnerability Index/domain-specific score and Neighborhood Deprivation Index in a census tract within New York City. The scatterplots include Spearman’s correlations between the overall Neighborhood Environmental Vulnerability Index and its domains with the Neighborhood Deprivation Index by the five boroughs in New York City.

**Supplemental Figure 2B. Correlations Between Neighborhood Environmental Vulnerability Overall Index and Domain-Specific Scores with Social Vulnerability Index by New York City Boroughs, 2015-2019**

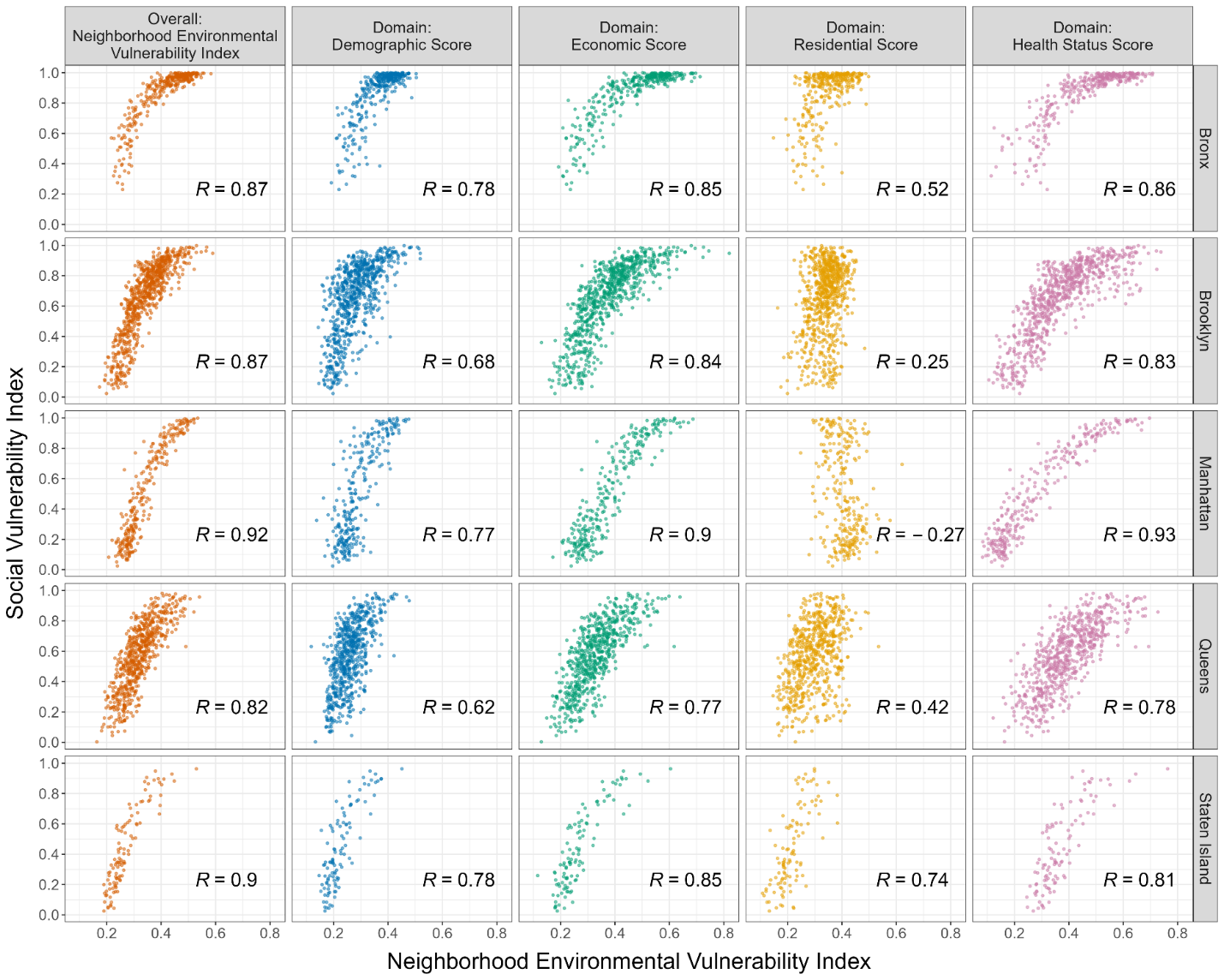

Each point represents the Neighborhood Environmental Vulnerability Index/domain-specific score and Neighborhood Deprivation Index in a census tract within New York City. The scatterplots include Spearman’s correlations between the overall Neighborhood Environmental Vulnerability Index and its domains with the Neighborhood Deprivation Index by the five boroughs in New York City.

**Supplemental Figure 3. Different Race and Ethnicity Categorizations: Distributions of NEVI, NEVI Demographic Domain Score, and Race and Ethnicity Subdomain Score**

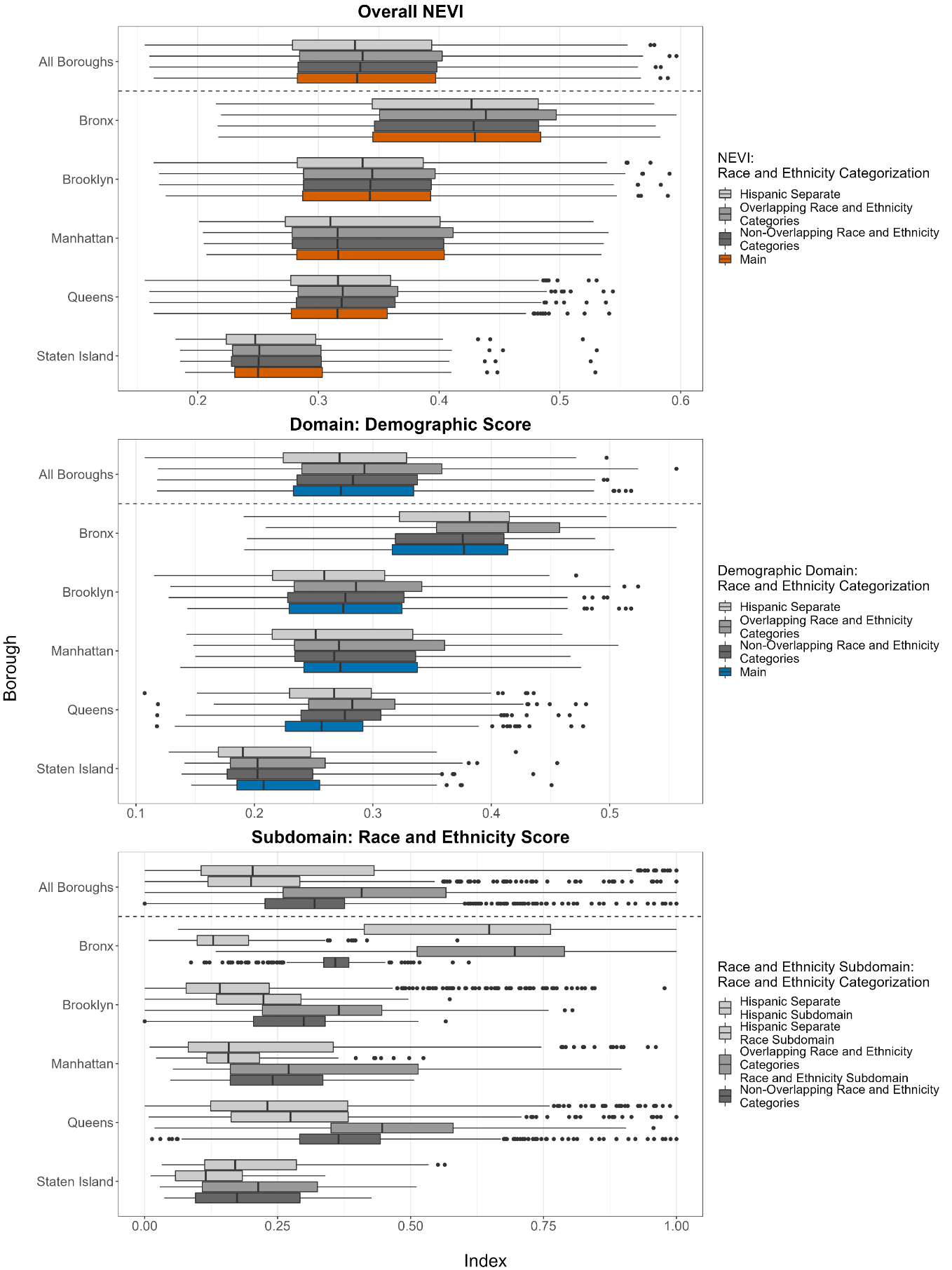

We did not include race and ethnicity in the NEVI (referred to as "Main” above). In this plot, we compared the distributions of NEVI, the NEVI demographic domain score, and the NEVI race and ethnicity subdomain scores with different categorizations of race: 1) overlapping race and ethnicity categories, 2) non-overlapping race and ethnicity categories, and 3) non-overlapping race and ethnicity categories with a separate Hispanic subdomain. The boxplots above display the median represented by the center line, first and third quartile represented by the lower and upper hinges, first or third quartile ± 1.5*IQR (interquartile range) represented by the lower and upper whiskers, and outliers beyond the whiskers represented by the dots.

**Supplemental Figure 4. NEVI including Race and Ethnicity Categories: Cluster Vulnerability Profiles for the Neighborhood Environmental Vulnerability Index across New York City, 2015-2019**

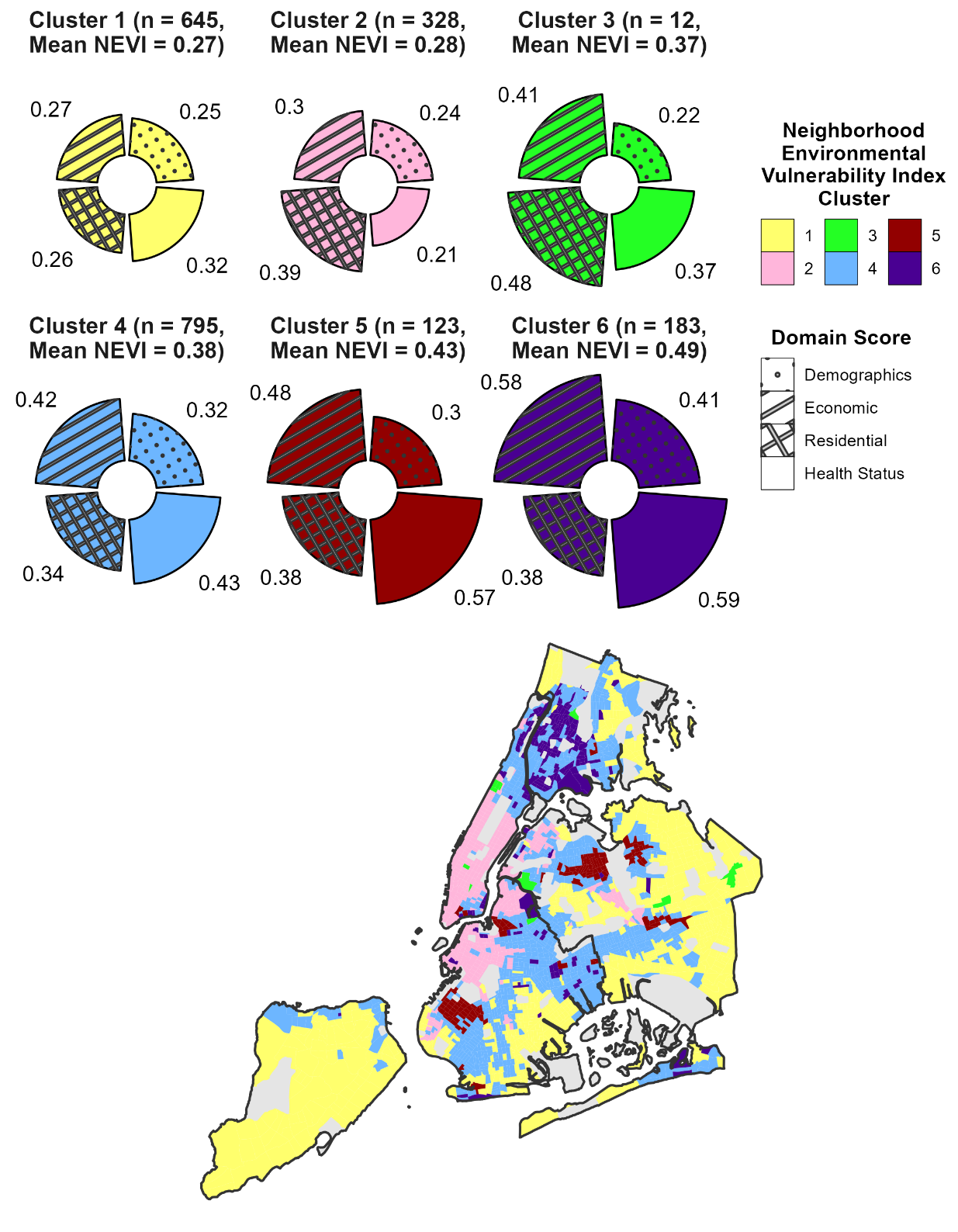

The circular barplots (top) shows the vulnerability profiles for the six clusters, with the different colors represent different clusters. The map (bottom) shows the distribution of the NEVI clusters across NYC, with areas that were excluded due to low population counts or missing features shown in dark gray. Data Sources: U.S. Census American Community Survey 2015-2019 5-Year Estimates and Centers for Disease Control and Prevention PLACES Project 2020 Release.

**Supplemental Text Calculating the Neighborhood Deprivation Index**

We briefly explained our calculation of the NDI in the main body of the paper and provide additional details here in the supplementary section. There are seven domains of deprivation used for NDI indicated by Messer et al: education, employment, housing, occupation, poverty (income), racial composition, and residential stability.(Messer et al., 2006) We incorporated an additional language domain to increase comparability between our version of the NDI and other existing indices describing social deprivation and vulnerability.(“CDC SVI Documentation 2018 | Place and Health | ATSDR,” 2021; Shmool et al., 2015) Within the eight final domains, we used 128 estimates drawn from the U.S. Census ACS to prepare 23 candidate features (Supplemental Table 2).

We first z-standardized all features and then reverse coded some features, such as percent population with at least Bachelor’s level education, to ensure that the final NDI had higher values indicating more deprivation. We also performed log transformation on median household income to ensure normal distribution. Next, we conducted successive principal component analyses (PCA) to identify a set of features that would maximize the variance explained with reduced dimensions and to develop an area-level NDI that reflects spatial heterogeneity across NYC boroughs. The successive PCA included an initial city-wide run, five spatially-stratified PCA iterations, a second city-wide run, and a final PCA feature extraction step with retained features. We started with 23 features for the first city-wide and five spatially stratified PCAs and applied varimax rotation to the loadings of the selected components (with eigenvalue > 1). For the first city-wide PCA, we retained features that 1) had a strong loading (greater than 0.25 in magnitude) for the first principal component and (Messer et al., 2006) 2) did not load prominently strong (greater than 0.4 in magnitude) on more than one component, so we could ensure that each feature only explain an specific domain.(Shmool et al., 2015) For the spatially stratified PCA, we built upon the previous two criteria and retain features that: 1) never loaded below 0.16 in magnitude any borough for the first component and (Messer et al., 2006) 2) loaded prominently strong (greater than 0.4 in magnitude) on only one component in two or more borough-level PCA solutions.(Shmool et al., 2015) After applying above criteria, we included 13 features and performed a second city-wide PCA on them. When the second city-wide PCA process was completed, we retained 7 final features: percent population with at least Bachelor’s level education, percent unemployed, adults in management or professional occupations, households in poverty (< 200% Federal Poverty Line), families with annual income less than $35,000, households with public assistance income, non-Hispanic non-White. Finally, we re-ran PCA with the 7 final NDI features and used the resulting score associated with the unrotated first principal component as the NDI. The features we retained in each step of our process are documented in Table 2.

**Supplemental Table 1. Summary Statistics Across Index and Domain-Specific Scores**

|  | **Borough** | **Mean** | **SD** | **Median** | **IQR** | **Min.** | **Max.** |
| --- | --- | --- | --- | --- | --- | --- | --- |
| **Index** |  |  |  |  |  |  |  |
| Neighborhood Environmental Vulnerability Index (NEVI) | **Overall** | 0.34 | 0.08 | 0.33 | 0.28-0.40 | 0.16 | 0.59 |
|  | Bronx | 0.41 | 0.09 | 0.43 | 0.35-0.48 | 0.22 | 0.58 |
|  | Brooklyn | 0.34 | 0.07 | 0.34 | 0.29-0.39 | 0.17 | 0.59 |
|  | Manhattan | 0.34 | 0.08 | 0.32 | 0.28-0.40 | 0.21 | 0.53 |
|  | Queens | 0.32 | 0.06 | 0.32 | 0.28-0.36 | 0.16 | 0.54 |
|  | Staten Island | 0.27 | 0.07 | 0.25 | 0.23-0.30 | 0.19 | 0.53 |
| Neighborhood Deprivation Index (NDI) | **Overall** | 0.00 | 1.00 | -0.01 | -0.67-0.58 | -2.16 | 3.61 |
|  | Bronx | 0.99 | 0.95 | 1.14 | 0.28-1.70 | -1.58 | 3.61 |
|  | Brooklyn | -0.01 | 0.91 | -0.01 | -0.58-0.48 | -2.02 | 3.41 |
|  | Manhattan | -0.76 | 1.16 | -1.20 | -1.77-0.17 | -2.16 | 2.44 |
|  | Queens | -0.10 | 0.61 | -0.07 | -0.53-0.27 | -1.85 | 2.08 |
|  | Staten Island | -0.36 | 0.80 | -0.61 | -0.88—0.09 | -1.35 | 3.36 |
| Social Vulnerability Index (SVI) | **Overall** | 0.63 | 0.26 | 0.67 | 0.43-0.85 | 0.00 | 1.00 |
|  | Bronx | 0.86 | 0.17 | 0.94 | 0.81-0.98 | 0.23 | 1.00 |
|  | Brooklyn | 0.65 | 0.24 | 0.70 | 0.50-0.84 | 0.02 | 1.00 |
|  | Manhattan | 0.50 | 0.31 | 0.45 | 0.20-0.81 | 0.02 | 1.00 |
|  | Queens | 0.57 | 0.22 | 0.59 | 0.42-0.74 | 0.00 | 0.98 |
|  | Staten Island | 0.44 | 0.26 | 0.36 | 0.23-0.62 | 0.03 | 0.96 |
| **NEVI Domain Score** |  |  |  |  |  |  |  |
| Demographics | **Overall** | 0.29 | 0.07 | 0.27 | 0.23-0.33 | 0.12 | 0.52 |
|  | Bronx | 0.36 | 0.07 | 0.38 | 0.32-0.41 | 0.19 | 0.50 |
|  | Brooklyn | 0.28 | 0.07 | 0.28 | 0.23-0.32 | 0.14 | 0.52 |
|  | Manhattan | 0.29 | 0.07 | 0.27 | 0.24-0.34 | 0.14 | 0.48 |
|  | Queens | 0.26 | 0.05 | 0.26 | 0.23-0.29 | 0.12 | 0.48 |
|  | Staten Island | 0.23 | 0.06 | 0.21 | 0.19-0.26 | 0.15 | 0.45 |
| Economic Indicators | **Overall** | 0.37 | 0.11 | 0.35 | 0.28-0.44 | 0.12 | 0.82 |
|  | Bronx | 0.46 | 0.12 | 0.48 | 0.37-0.57 | 0.19 | 0.71 |
|  | Brooklyn | 0.38 | 0.10 | 0.37 | 0.30-0.44 | 0.13 | 0.82 |
|  | Manhattan | 0.38 | 0.11 | 0.36 | 0.29-0.47 | 0.17 | 0.69 |
|  | Queens | 0.33 | 0.08 | 0.32 | 0.27-0.38 | 0.13 | 0.64 |
|  | Staten Island | 0.27 | 0.09 | 0.25 | 0.22-0.30 | 0.12 | 0.60 |
| Residential Characteristics | **Overall** | 0.33 | 0.07 | 0.33 | 0.28-0.38 | 0.11 | 0.62 |
|  | Bronx | 0.35 | 0.07 | 0.34 | 0.29-0.40 | 0.15 | 0.50 |
|  | Brooklyn | 0.34 | 0.05 | 0.34 | 0.31-0.37 | 0.16 | 0.48 |
|  | Manhattan | 0.41 | 0.05 | 0.41 | 0.37-0.44 | 0.25 | 0.62 |
|  | Queens | 0.30 | 0.07 | 0.30 | 0.25-0.35 | 0.15 | 0.53 |
|  | Staten Island | 0.22 | 0.06 | 0.21 | 0.18-0.25 | 0.11 | 0.38 |
| Health Status | **Overall** | 0.38 | 0.13 | 0.38 | 0.29-0.48 | 0.08 | 0.76 |
|  | Bronx | 0.46 | 0.13 | 0.49 | 0.35-0.56 | 0.11 | 0.71 |
|  | Brooklyn | 0.37 | 0.12 | 0.36 | 0.28-0.45 | 0.11 | 0.74 |
|  | Manhattan | 0.29 | 0.16 | 0.24 | 0.16-0.42 | 0.08 | 0.70 |
|  | Queens | 0.39 | 0.10 | 0.39 | 0.32-0.47 | 0.12 | 0.73 |
|  | Staten Island | 0.37 | 0.09 | 0.35 | 0.31-0.41 | 0.25 | 0.76 |

Acronyms in the table: SD = standard deviation, IQR = interquartile range

**Supplemental Table 2. Features Selected in the Neighborhood Deprivation Index**

| **Candidate Features (23 features)** | **Retained Features** | | |
| --- | --- | --- | --- |
|  | **Step 1:  City-wide PCA (9 features)** | **Step 2:  Borough-stratified PCA (10 features)** | **Step 3:  Second City-wide PCA (7 features)** |
| **Education**, among adults aged > 25 | | | |
| • With less than high school education (%) |  | **✓** |  |
| • With at least Bachelor's-level education (%) |  | **✓** | **✓** |
| **Employment**, among adult labor force aged 20-64 | | | |
| • Unemployed (%) | **✓** |  | **✓** |
| • Male in labor force (%) |  |  |  |
| • Female in labor force (%) |  |  |  |
| **Housing** | | | |
| • Renter-occupied, among occupied units (%) | **✓** | **✓** |  |
| • Vacancy, among total housing units (%) |  |  |  |
| • Crowded (>1 occupant per room, among occupied housing units) (%) | **✓** | **✓** |  |
| **Occupation**, among full-time, year-round civilian employed population | | | |
| • Adults in management or professional occupations (%) |  | **✓** | **✓** |
| **Income (Poverty)** | | | |
| • Households in poverty, <200% Federal Poverty Line (%) | **✓** | **✓** | **✓** |
| • Families w/ annual income <$35,000, 2019 inflation-adjusted (%) | **✓** | **✓** | **✓** |
| • Female-led households (%) |  |  |  |
| • Households with public assistance income (%) | **✓** |  | **✓** |
| • Households with food stamp benefits (%)* |  |  |  |
| • Household Income (median)* | **✓** | **✓** |  |
| • Renter- or owner- housing costs in excess of 30% household income (%)* |  |  |  |
| **Racial composition** | | | |
| • Black or African American, non-Hispanic (%) |  |  |  |
| • Not non-Hispanic White (%)** | **✓** |  | **✓** |
| • Hispanic or Latino (%) |  | **✓** |  |
| **Residential Stability** | | | |
| • Living in the same house one year ago (%) |  |  |  |
| • Foreign-born (%) |  |  |  |
| • Not a U.S. citizen (%) |  |  |  |
| **Language** | | | |
| • Speak English less than “very well” (%)*** | **✓** | **✓** |  |

***Data source****: U.S. Census American Community Survey (ACS) 2015-2019 5-year estimates*

** In the past 12 months*

*** Calculated as 1 minus the percent of Non-Hispanic White population*

**** Among population 5 years or older who speak a language other than English at home*

**Supplemental Table 3. Neighborhood Deprivation Index, Borough-Specific and City-Wide feature Loadings on the First Principal Component**

|  | **Bronx** | **Brooklyn** | **Manhattan** | **Queens** | **Staten Island** | **City-wide Index** | **Loading Difference** |
| --- | --- | --- | --- | --- | --- | --- | --- |
| **% B.S./B.A. or higher** | 0.89 | 0.87 | 0.97 | 0.84 | 0.77 | 0.85 | 0.2 |
| **% Unemployment** | 0.62 | 0.53 | 0.59 | 0.44 | 0.24 | 0.61 | 0.4 |
| **% Management Occupation** | 0.85 | 0.82 | 0.92 | 0.82 | 0.71 | 0.82 | 0.2 |
| **% Households in Poverty** | 0.89 | 0.79 | 0.91 | 0.65 | 0.93 | 0.82 | 0.3 |
| **% Households with annual income < $35,000** | 0.89 | 0.86 | 0.95 | 0.58 | 0.90 | 0.84 | 0.4 |
| **% Households receiving public assistance** | 0.77 | 0.74 | 0.85 | 0.60 | 0.90 | 0.77 | 0.3 |
| **% Non-Hispanic non-White** | 0.79 | 0.63 | 0.94 | 0.75 | 0.86 | 0.74 | 0.3 |
| **% Variance Explained** | 67.3 | 57.0 | 79.1 | 46.5 | 62.3 | 61.3 |  |

**Supplemental Table 4. Subdomain Summary Statistics of NEVI Clusters**

| **Subdomain  (Weight)** | **Cluster Median (IQR)** | | | | | |
| --- | --- | --- | --- | --- | --- | --- |
|  | **1  (n = 435, 20.85%)** | **2 (n = 331, 15.87%)** | **3 (n = 903, 43.29%)** | **4 (n = 5,**  **0.24%)** | **5 (n = 241, 11.55%)** | **6 (n = 171, 8.20%)** |
| Overall NEVI | 0.26 (0.23-0.28) | 0.28 (0.26-0.30) | 0.35 (0.32-0.38) | 0.44 (0.41-0.45) | 0.46 (0.42-0.48) | 0.47 (0.43-0.50) |
| Age  (1/32) | 0.45 (0.40-0.49) | 0.30 (0.21-0.39) | 0.39 (0.34-0.44) | 0.68 (0.55-0.77) | 0.41 (0.37-0.45) | 0.44 (0.39-0.49) |
| Female-led household (1/32) | 0.12 (0.07-0.22) | 0.06 (0.03-0.10) | 0.22 (0.14-0.34) | 0.02 (0.00-0.08) | 0.32 (0.17-0.50) | 0.54 (0.44-0.67) |
| Immigration  (1/32) | 0.22 (0.15-0.27) | 0.24 (0.19-0.30) | 0.32 (0.25-0.41) | 0.58 (0.38-0.64) | 0.48 (0.4-0.6) | 0.24 (0.20-0.32) |
| Disability (1/32) | 0.15 (0.11-0.18) | 0.11 (0.08-0.15) | 0.15 (0.11-0.20) | 0.57 (0.44-0.81) | 0.17 (0.11-0.23) | 0.27 (0.22-0.32) |
| Single Parent Households  (1/32) | 0.21 (0.13-0.39) | 0.17 (0.11-0.30) | 0.39 (0.26-0.56) | 0.15 (0.15-0.21) | 0.49 (0.27-0.66) | 0.75 (0.66-0.84) |
| Mobility (1/32) | 0.14 (0.11-0.18) | 0.28 (0.25-0.32) | 0.22 (0.17-0.27) | 0.20 (0.17-0.24) | 0.27 (0.21-0.31) | 0.25 (0.22-0.29) |
| Social Isolation (1/32) | 0.23 (0.18-0.28) | 0.44 (0.35-0.54) | 0.27 (0.18-0.36) | 0.54 (0.49-0.70) | 0.23 (0.16-0.29) | 0.35 (0.29-0.42) |
| Income and poverty  (1/24) | 0.41 (0.37-0.44) | 0.36 (0.28-0.43) | 0.53 (0.48-0.59) | 0.79 (0.70-0.85) | 0.67 (0.59-0.73) | 0.76 (0.70-0.81) |
| Occupation (1/24) | 0.40 (0.31-0.47) | 0.14 (0.08-0.23) | 0.51 (0.43-0.59) | 0.65 (0.58-0.76) | 0.69 (0.61-0.75) | 0.64 (0.57-0.71) |
| Income inequality  (1/24) | 0.30 (0.24-0.37) | 0.44 (0.35-0.54) | 0.38 (0.30-0.46) | 0.56 (0.47-0.58) | 0.41 (0.34-0.48) | 0.49 (0.42-0.59) |
| Unemployment (1/24) | 0.12 (0.08-0.17) | 0.10 (0.07-0.14) | 0.16 (0.11-0.22) | 0.24 (0.07-0.30) | 0.18 (0.10-0.28) | 0.31 (0.19-0.42) |
| Education (1/24) | 0.16 (0.12-0.22) | 0.08 (0.03-0.13) | 0.28 (0.21-0.37) | 0.22 (0.15-0.23) | 0.53 (0.45-0.60) | 0.46 (0.39-0.57) |
| Vehicle availability  (1/24) | 0.10 (0.05-0.17) | 0.66 (0.54-0.75) | 0.37 (0.23-0.54) | 0.64 (0.59-0.67) | 0.61 (0.52-0.70) | 0.65 (0.53-0.76) |
| Population Density (1/28) | 0.08 (0.05-0.12) | 0.24 (0.16-0.36) | 0.17 (0.12-0.25) | 0.29 (0.06-0.34) | 0.31 (0.22-0.44) | 0.23 (0.16-0.33) |
| Group Quarters (1/28) | 0.00 (0.00-0.01) | 0.00 (0.00-0.03) | 0.00 (0.00-0.01) | 0.00 (0-0) | 0.00 (0.00-0.02) | 0.01 (0.00-0.03) |
| Occupants per room (1/28) | 0.07 (0.03-0.12) | 0.09 (0.05-0.12) | 0.17 (0.11-0.25) | 0.19 (0.09-0.31) | 0.36 (0.27-0.46) | 0.17 (0.11-0.23) |
| Age of housing structure (1/28) | 0.84 (0.72-0.97) | 1.00 (0.7-1.0) | 0.97 (0.81-1.00) | 1.00 (0.92-1.00) | 1.00 (0.82-1.00) | 0.74 (0.58-0.82) |
| Units in housing structure (1/28) | 0.49 (0.32-0.57) | 0.87 (0.73-0.94) | 0.68 (0.57-0.81) | 0.93 (0.91-0.98) | 0.82 (0.66-0.90) | 0.89 (0.80-0.95) |
| Changing residence (1/28) | 0.15 (0.10-0.22) | 0.39 (0.3-0.5) | 0.20 (0.13-0.29) | 0.30 (0.24-0.30) | 0.19 (0.13-0.25) | 0.15 (0.09-0.20) |
| Housing vacancy (1/28) | 0.11 (0.07-0.16) | 0.20 (0.13-0.30) | 0.13 (0.09-0.18) | 0.10 (0.09-0.13) | 0.11 (0.06-0.15) | 0.09 (0.05-0.14) |
| Unhealthy behaviors (1/16) | 0.36 (0.28-0.44) | 0.28 (0.22-0.34) | 0.47 (0.41-0.53) | 0.29 (0.29-0.47) | 0.61 (0.55-0.66) | 0.67 (0.61-0.72) |
| Health outcomes (1/16) | 0.31 (0.28-0.34) | 0.18 (0.15-0.23) | 0.33 (0.29-0.36) | 0.57 (0.54-0.62) | 0.37 (0.32-0.42) | 0.47 (0.41-0.51) |
| Prevention practices (1/16) | 0.32 (0.26-0.38) | 0.27 (0.18-0.35) | 0.49 (0.40-0.57) | 0.42 (0.41-0.46) | 0.69 (0.62-0.77) | 0.61 (0.55-0.67) |
| Health insurance access (1/16) | 0.19 (0.15-0.24) | 0.10 (0.05-0.17) | 0.34 (0.26-0.42) | 0.17 (0.17-0.32) | 0.57 (0.51-0.64) | 0.54 (0.47-0.62) |
